# Tangent Approximation Formula to Determine Height Loss in Scoliosis Patients by a new Trigonometric Model

**DOI:** 10.1101/2025.03.22.25324448

**Authors:** Tsz-Hin Tang, Nam Lau, Tsz-Ping Lam, Adam Yiu-Chung Lau, Alec Lik-Hang Hung, Winnie C.W. Chu, Jack Chun-Yiu Cheng, Nelson L.S. Tang

## Abstract

**Background:** The problem of predicting loss in body height due to scoliosis especially in adolescent idiopathic scoliosis (AIS) has been investigated for over 50 years. The corrected height is required to make appropriate lung function and other anthropometry assessments. Most existing formulae estimate height loss through different regression models.

**Method:** A new trigonometric model was developed based on the Cobb angle (α) and the apical vertebral translation (AVT) in the coronal plane. By using this model, a formula was derived using the tangent of the triangle to estimate height loss in Scoliosis.

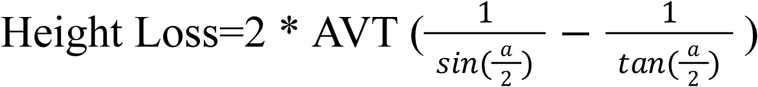

As measurement of AVT is not commonly performed, a mean value was obtained from patients series reported by Şarlak et al. It was found that AVT is affected by the Cobb angle, which was significantly different between patients with Cobb angle >= 50 and < 50.

**Results:** Validation of this new Tangent formula was carried out with two datasets, one from Tyrakowski et al 2014, and another from 58 operated Scoliosis patients in a local hospital. The mean bias was the lowest with this novel tangent formula compared to Bjure, Kono, Stokes formulae. A paired t-test confirmed that these differences were significantly better with the tangent formula. The trigonometric approach has the advantage that it is valid over a larger range of Cobb angle as most of the regression models cannot be extrapolated outside the range of available data points.

The Tangent formula is made available at the website for easy access and height loss can be estimated by inputting the Cobb angle with or without the AVT: https://tangentformulae.scoliosisdevices.com/

**Conclusion:** The results indicate the use of the Tangent formula with simple measurements and trigonometric calculation yield results comparable with other formulae used in the past to estimate height loss. The application of this new formula can prove useful in clinical settings to allow more accurate testing and better care for patients.

## Introduction

The motivation to estimate the loss in body height due to curvature of scoliosis has been around for 50 years since Bjure et al^1^, Johnson et al^2^ and Lindh et al^3^. These studies aimed to estimate lung function parameters using corrected body height. This extended to growth chart, body mass index and estimation of residual growth^4^. Gain in height after curve correction surgery is a common phenomenon in both AIS^5, 6^ and adult spinal deformity^7^. Alternative measurements such as arm span were also suggested to measure body height in scoliosis patients^8^. Body height also has an important contribution of self-image of patients^9^.

The current common method to estimate height loss is through regression, this could be a linear function of the Cobb angle^10^ or a power function of Cobb angle^3, 11, 12^ (see appendix). The regression models are limited by the range of Cobb angle values in the dataset and have limited ability to extrapolate. Şarlak et al made the first attempt to determine height loss by using a circle to model the scoliosis curve^9, 13^. However, the method required input of many measurements. In addition to the Cobb angle itself, making it less applicable for routine clinical use. An ideal method which is both unrestricted by the dataset and simple to use is yet to be found.

We propose a novel trigonometric model to approximate the scoliosis curve by isosceles triangles and height loss can be solved mathematically by trigonometric functions using the Cobb angle and the apical vertebral translation (AVT) in the coronal plane.

## Materials and Methods

### Trigonometry model underlying the tangent formulae

The curve in scoliosis is measured in terms of Cobb angle. The angle between the upper and lower endplates of the vertebrae in the scoliosis curve is the Cobb angle.

In figure 1A, it is shown that the scoliosis spine could be modelled by two isosceles triangles (ΔABD and ΔBCD) where the curve of the scoliotic vertebrae forms the two congruent sides of ΔBCD (Figure 1A).

**Figure 1.**
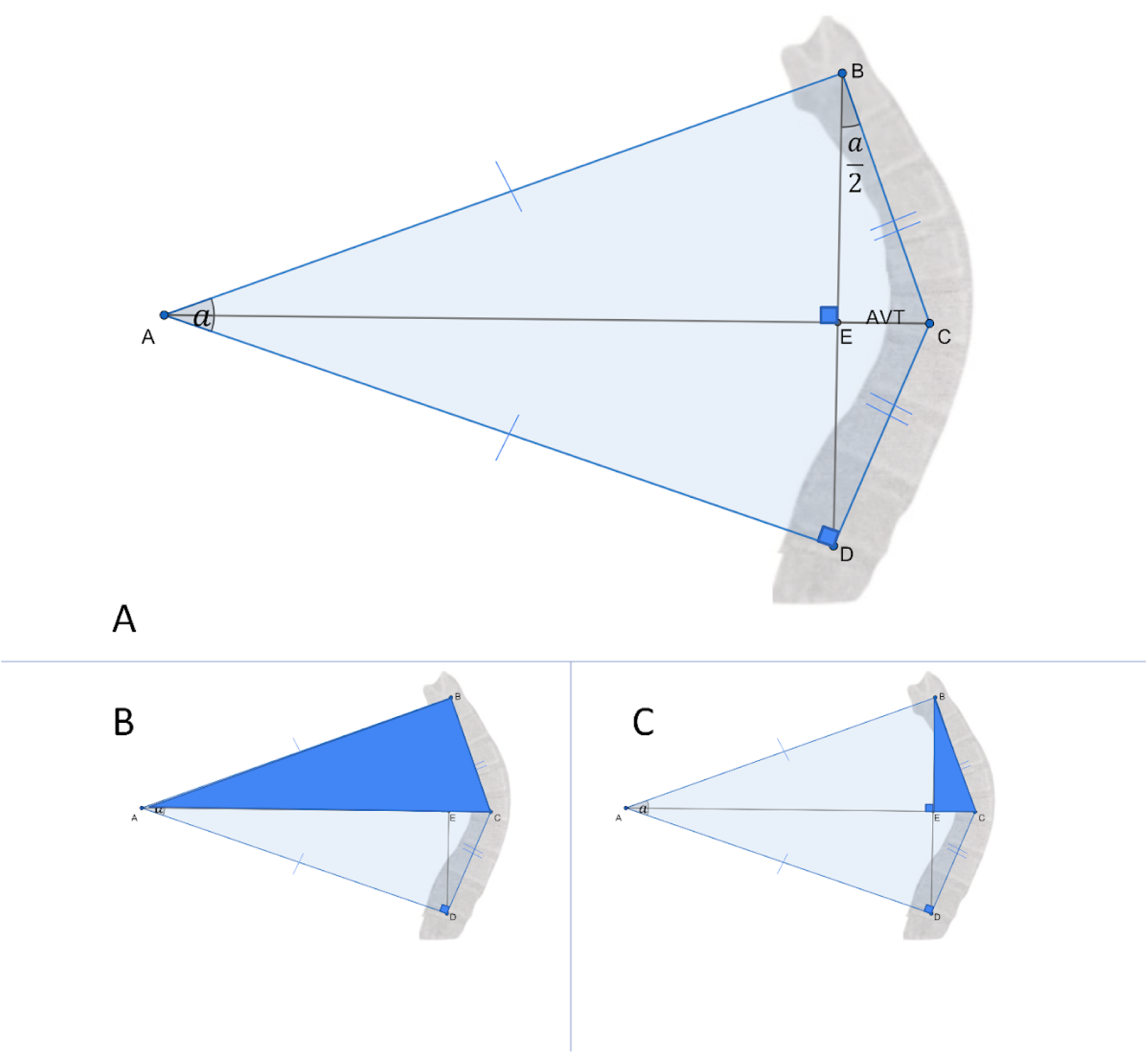
Principle of the new Trigonometric model of the scoliotic segment of spine. α is the Cobb angle. Segment EC represents the apical vertebral translation (AVT). Figure 1A shows the overall view. Figure 1B and 1C show a series of two different right angle triangle which form the basis of the trigonometric model. Segments BE + ED constitute the observed height (before operation) and segments BC + CD constitute the true height (after operation). The difference between these 2 lines is the height loss.

ΔABE and its identical mirror image ΔADE are right angle triangles. The ∠ABC is a right-angle (Figure 1B). Under this scheme, segments BE and ED represent pre-operative height. Both congruent sides added (segments BC + CD) together post-operative height / corrected height. This difference of BC+CD and BE+ED can be determined by the length difference of one of the right triangle (i.e. the difference between segment BC and BE in Figure 1C) and multiply by 2.

The dimensions for segments BC and BE can be derived from the ΔBEC (Figure 1C). ∠CBE is equal to ∠BAC, i.e half of Cobb angle. The segment CE is known clinically as AVT. With AVT value, the difference between the hypotenuse segments BC and BE can be determined by trigonometry functions. Therefore the New Tangent Formula can be derived as:

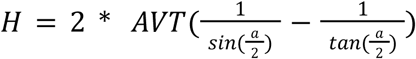

Where *a* = Cobb Angle

If AVT is not measured, a mean value is assigned. A more accurate estimate of height loss can be obtained if AVT is measured from patient’s radiograph.

### Mean values of AVT in Şarlak et al

Şarlak et al measured both AVT and Cobb angles from calibrated pre- and post-operative anteroposterior (AP) radiographs of thirty-six patients who underwent posterior instrumentation and fusion^13^. This allowed mean AVT values to be determined in two patient group of Cobb angle below 50 or above 50 degree.

#### Using ½ of Cobb angle and approximated AVT to determine height loss

Based on the 2 Cobb angle subgroups, the mean AVT was used as an approximation AVT together with (Cobb angle)/2 to predict height loss in AIS patients. The estimated height loss were validated with two additional datasets. The first dataset were 30 patients reported in Tyrakowski et al^14^. The second dataset was a retrospective study of 58 scoliosis patients who participated in an ongoing bone mineralization study and received surgery in a local hospital in Hong Kong over a period of 4 years. Standing body height was measured without shoes with the head in the Frankfort horizontal plane against a wall-mounted stadiometer (at 0.1cm accuracy). Patients with congenital scoliosis, neuromuscular diseases, skeletal dysplasia, endocrine diseases or history of hormonal treatment were excluded. Most patients (21) were classified as Lenke type 1, 11 were Lenke type 2, 20 were type 3. 3 or less patients of other Lenke types were included. The mean (SD) age of patients was 16 (2.4) years. In this dataset, the heights of patients were measured before and at least one month after operation. Comparison with other formulae of height loss was made to the observed height loss to determine the absolute difference, limits of agreement (LOA), root mean square error (RMSE), and bias of estimates over the range of Cobb angles.

### Statistical analysis

Various estimating formulae were compared to the observed height loss (ground truth) by standard method-comparison parameters^15^. Mean bias of each formula was obtained by subtracting the observed height loss from estimated height loss and a positive bias represents over-estimation by the prediction formula. Limits of agreement were calculated by mean bias ± 1.96 x SD of bias and were shown in Bland-Altman plots as dotted red lines. Bias was compared between the new tangent formula against other formulae in a paired t test. Statistics were performed in MedCalc version 23 or excel.

## Results

### AVT and Cobb angle

Preoperational AVT in mm was plotted for the 2 subgroups of patients in Figure 2 using data reported by Şarlak et al^13^. Patients were divided into groups of Cobb angle <50 degree or ⩾50 degrees. The mean (SD) of AVT was 3.97 cm (1.46) for 24 patients with lower Cobb angle. Mean (SD) AVT was 5.37 cm (1.74) for Cobb angle ⩾50 degree (N=12). The difference in the AVT values of the 2 groups was statistically significant (p < 0.05). And rounded values of two means (4.0 and 5.4 cm) were used in the tangent formula.

**Figure 2.**
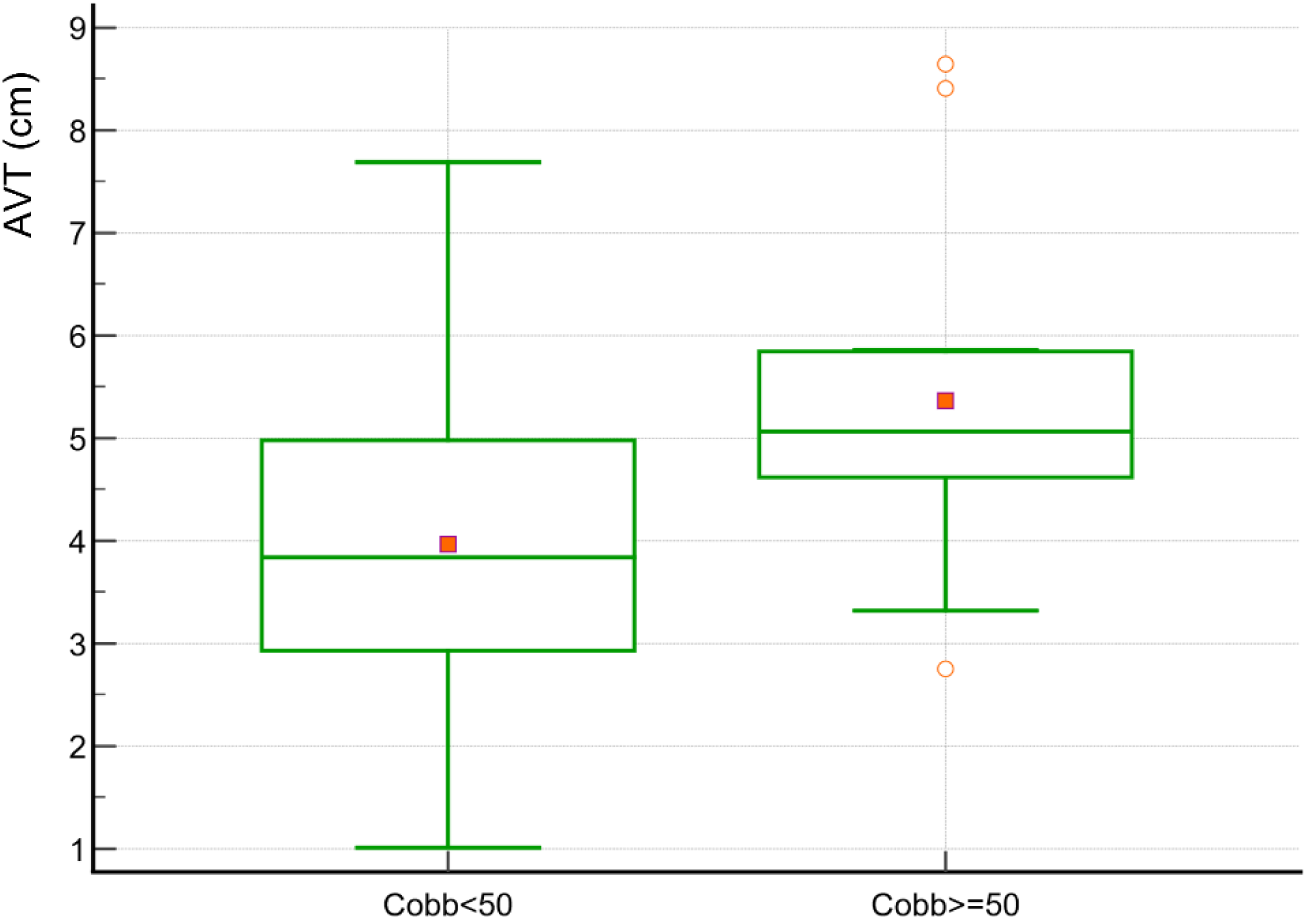
Distribution of AVT between patients with Cobb angle <50 degree and ⩾50 degree. Different between 2 groups was statistically significant (p<0.05)

### Determination of loss of height by the tangent formulae using mean AVT

Validation by correlation analysis between estimated and measured height loss with 30 patients reported by Tyrakowski et al. Predicted height loss was estimated by a series of formulae (shown in Appendix) in addition to the tangent formulae using mean AVT. The maximal Cobb angle of the major curve was used.

Patients in the dataset from Tyrakowski et al had complete data of Cobb angles of 3 vertebral segments together with measured height loss^14^. Predicted values of the three other formulae were available from Table 1 in Tyrakowski et al. Correlation coefficients between observed and predicted loss in height were comparable across models, including Stoke formula (R=0.94), the new Tangent formula (R=0.94), and Bjure formula (R=0.94) while it was slightly lower with the Kono formulae (R=0.92). Due to the small difference between correlation coefficients, standard method comparison statistics are used to compare the various formulae.

**Table 1.** Comparison of mean bias and LOA of height loss prediction formulae in Tyrakowski et al dataset (N=30). Prediction formulae are list by alphabetical order. Values are shown in cm. The new tangent formula showed best performance in 3 out of 4 parameters. **#** indicate formulae with a significant bias (ie the mean bias was not zero). The best-performing formula is shown in Bold font for each comparison parameter.

|  | Mean bias | 95% CI of mean bias | LOA | Median Absolute % error |
| --- | --- | --- | --- | --- |
| Bjure | 0.55 | 0.38 to 0.71 # | -0.33 to 1.42 | 43 % |
| Kono | -1.02 | -1.29 to -0.75 # | -2.44 to 0.40 | 54 % |
| new Tangent | **0.15** | **-0.03 to 0.32** | -0.77 to 1.06 | **21 %** |
| Stokes | 0.30 | 0.17 to 0.44 # | **-0.18 to 1.42** | **21 %** |

Figure 3 shows the Bland-Altman plot of this dataset. The new Tangent formula (Figure 3) showed the lowest bias. All other prediction formulae had a statistically significant mean bias.

**Figure 3.**
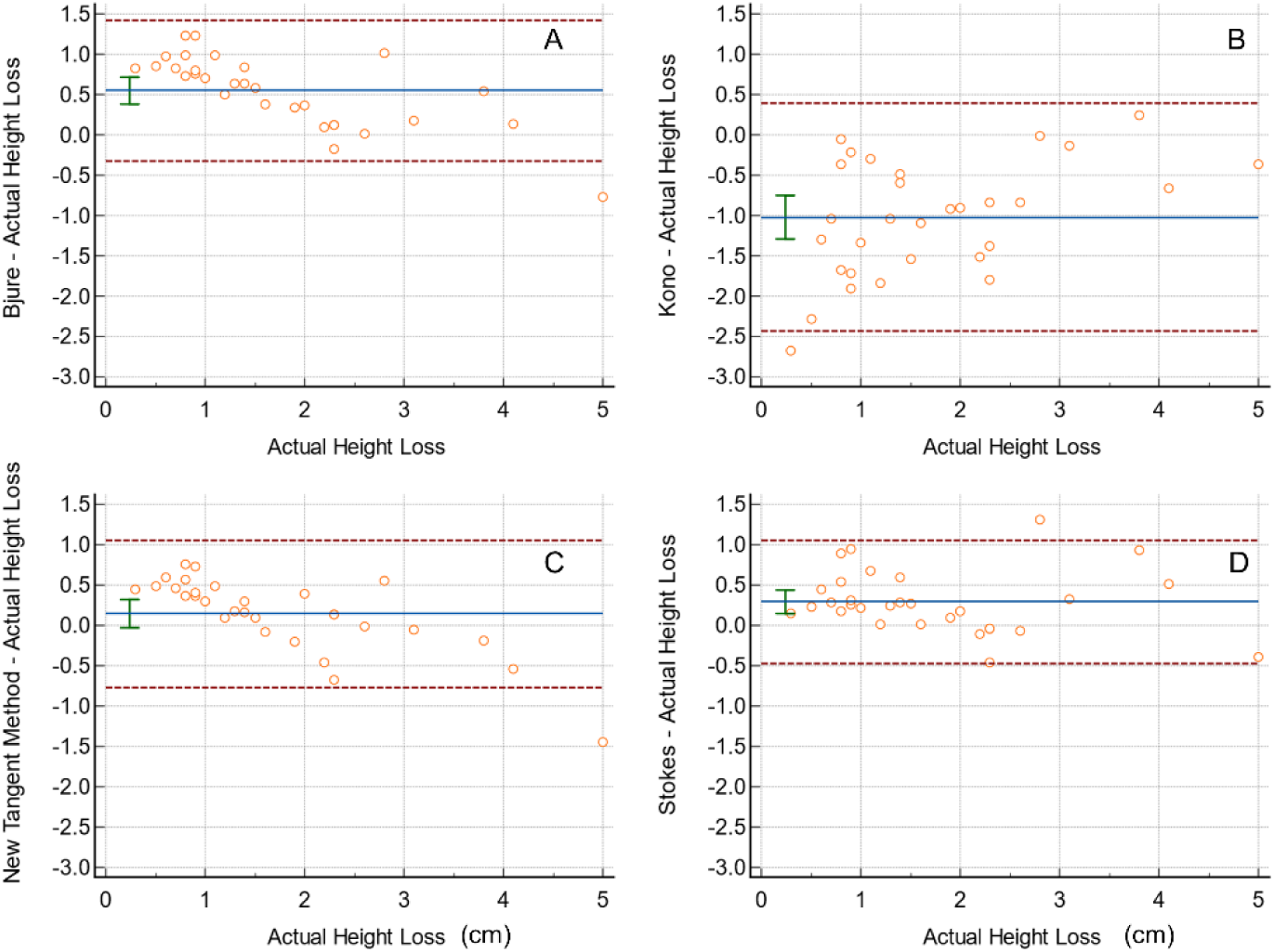
Bland-Altman plot of Tyrakowski, et al dataset (N=30). Four estimation formulae are shown and arranged by alphabetical order. A: Bjure formulae, B: Kono formula, C: new Tangent formula and D: Stokes formula. Mean bias is shown as blue line and LOA are dotted red lines. 95% Confidence intervals of mean bias are shown as error bars.

Results of the bias and LOA among prediction formulae are shown in Table 1. The new Tangent formula had a distinct advantage with the least bias. All three other prediction formulae had a significant mean bias with the range of 95% CI of mean bias outside of 0 (Error bars in Figure 3). Similarly, the absolute percentage error was the lowest in the new Tangent formula (21%) compared to others (Bjure 43% and Stokes 44%).

Validation by correlation analysis between estimated and measured height loss among patients operated in a local hospital

58 AIS patients who have undergone surgical corrections (posterior spinal fusion with pedicle screw instrumentation system) in a local hospital were analysed for bias of estimated height loss as calculated by various formulae. The mean ± standard deviation of pre-operative and post-operative body height were 156.3 ± 6.3 cm and 159.8 ± 6.2 cm, respectively. The mean maximal Cobb angle was 67.5 ± 12.8 before the operation (range: 45.0-103.0). The mean height loss was 3.5 ± 1.6 cm (range: 0.0-8.0cm).

Figure 4A shows the dot and line plot comparing the actual height loss and predicted height loss by the new tangent formula. The mean predicted height loss was 3.3 ± 0.7 cm. The mean predicted height loss matched well with observed height loss and were not different in a paired t test. Compared to the Kono formula in Figure 4B, the new Tangent formula provides height loss estimates with less bias.

**Figure 4.**
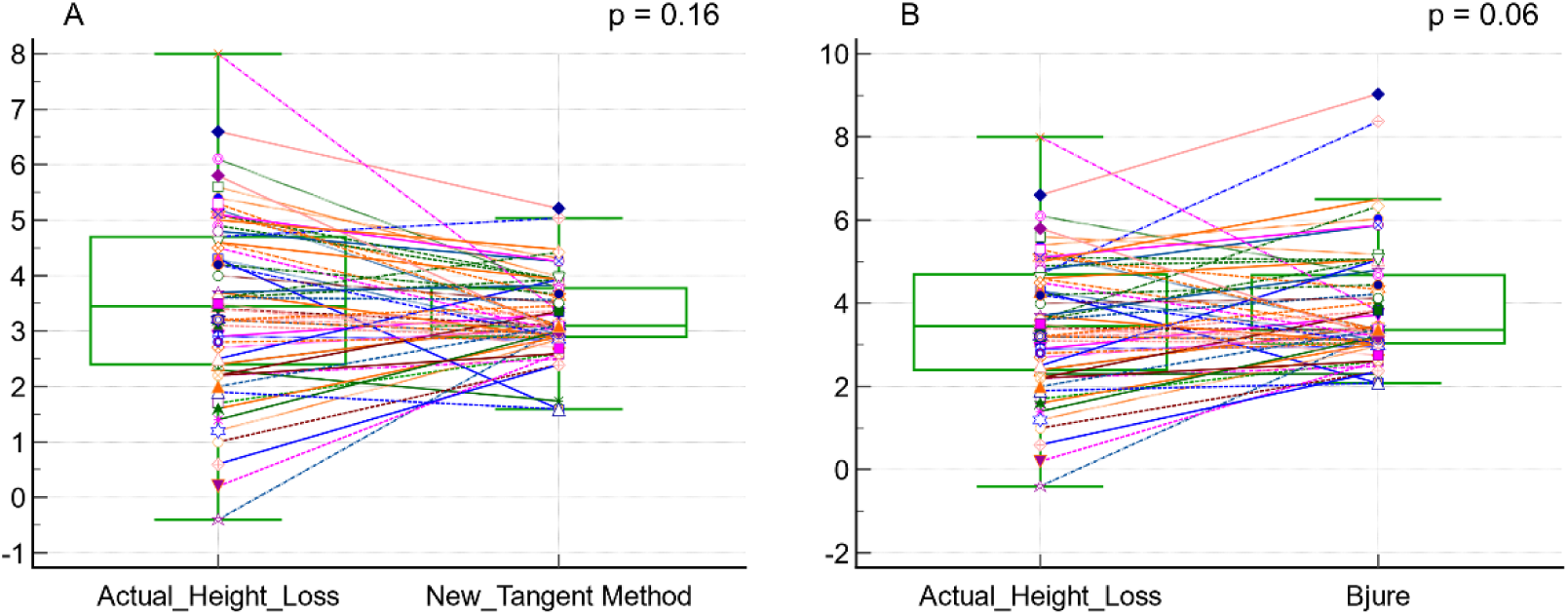
Dot and line plot of actual height loss and predicted height loss by the new tangent formula Figure 4A Dot and line plot compares actual height loss with predicted values by the new tangent formula. Paired t test was not significant. Figure 4B Dot and line plot compares actual height loss with predicted values by Bjure formula. Paired t test P value ¬ 0.06.

Using the Bland-Altman plots, the mean bias and LOA are shown in Figure 5. Both Kono and Stokes had a statistically significant mean bias. Furthermore, the new Tangent formulae also showed the best performance with the least absolute % error in Table 2 (22% vs 26%, 31% and 34%). Paired t test was performed to compare the bias value of the new Tangent formula against each of the 3 other formulae. All comparisons were statistically significant supporting that the new Tangent formula performs better compared to other formulae. However, a negative trend for the bias across the range of height loss for all 4 formulae.

**Figure 5.**
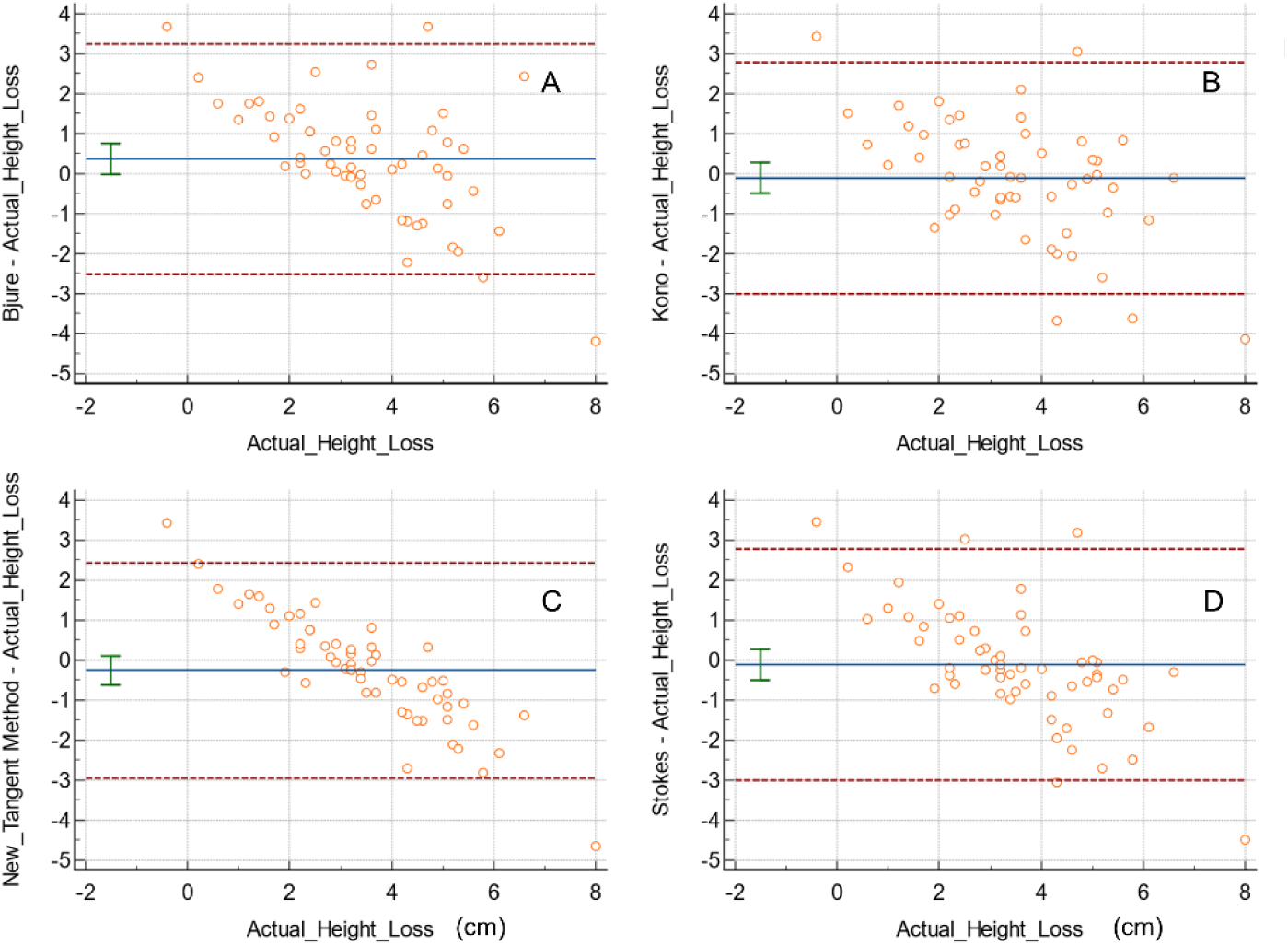
Bland-Altman plot of local patients who received surgical correction of scoliosis (N=58). Four estimation formulae are shown and arranged by alphabetical order. A: Bjure formulae, B: Kono formula, C: new Tangent formula and D: Stokes formula. 95% Confidence intervals of mean bias are shown as error bars.

**Table 2.** Comparison of bias and LOA of of height loss prediction formulae in local patients (N=58). Prediction formulae are list by alphabetical order. Values are shown in cm. The best-performing formula is/are shown in Bold font for each comparison parameter. RMSE: root mean square error

|  | Mean bias | 95% CI of mean bias | LOA | Median Absoluite % error | RMS E |
| --- | --- | --- | --- | --- | --- |
| Bjure | 0.37 | -0.01 to 0.76 | -2.50 to 3.25 | 26% | 1.50 |
| Kono | **-0.10** | -0.49 to 0.28 | -2.99 to 2.78 | 24% | 1.46 |
| new Tangent | -0.26 | -0.62 to 0.11 | **-2.96 to 2.44** | **22%** | **1.39** |
| Stokes | **-0.11** | -0.50 to 0.27 | -3.00 to 2.77 | 25% | 1.46 |

The issue of over-estimation of predicted height loss with some formulae has been recognised. Figure 6 shows the extent of over-estimation (a positive Y-axis value) against the maximal Cobb angle (Max Cobb angle) of the local hospital case series. The height loss in two patients with the top Cobb angle range were over-estimated by Bjure formula (arrows in Figure 6A) while Stokes and Kono formulae overestimated one(Figure 6). In comparison, there was no significant overestimation for the two using the new Tangent formula.

**Figure 6.**
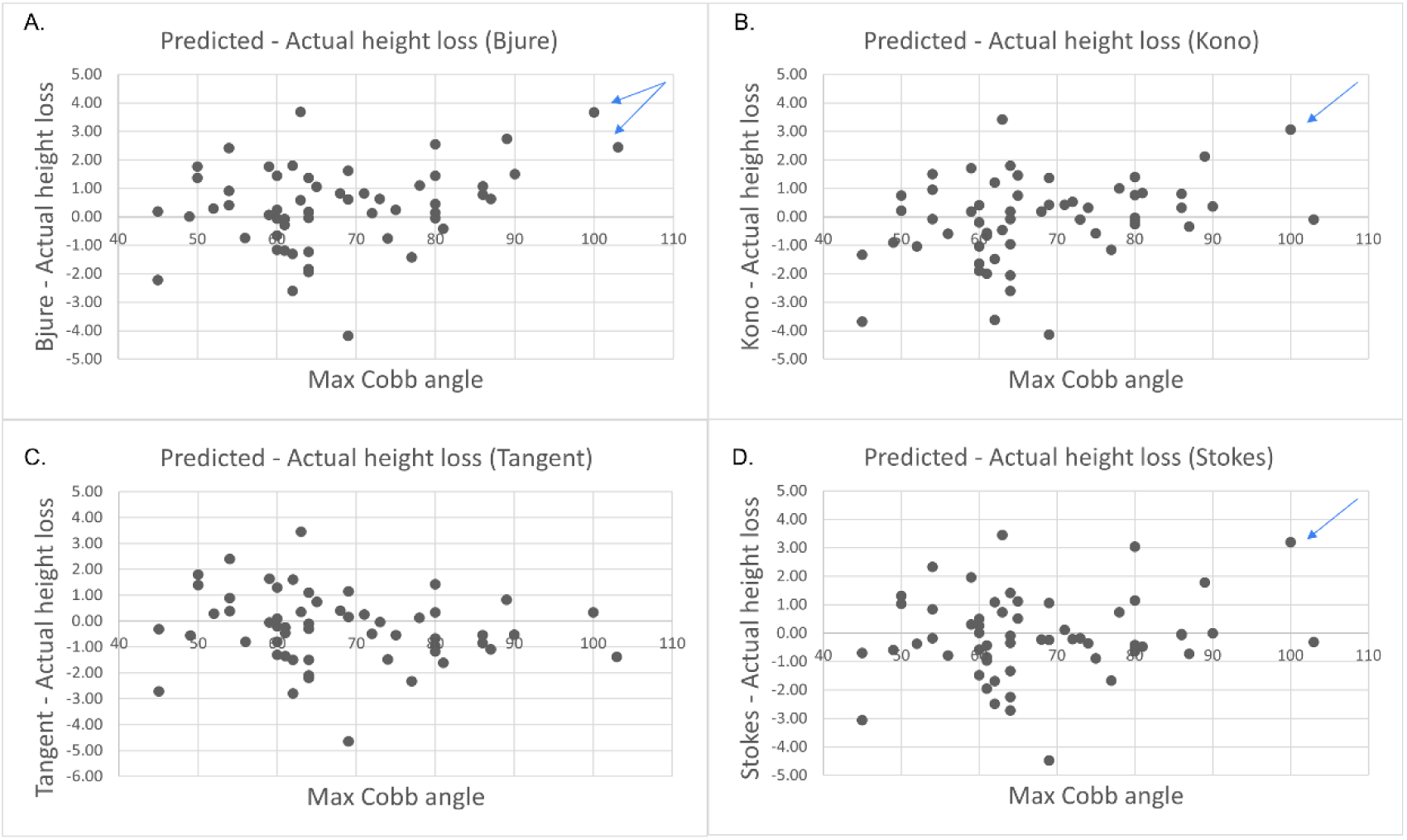
These graphs shows the difference between predicted height loss to actual height loss against maximal Cobb angle while using various prediction formulae. The local hospital data were used (N=58). Four estimation formulae are shown and arranged by alphabetical order. A: Bjure formulae, B: Kono formula, C: new Tangent formula and D: Stokes formula. Arrows point to those points that were over-estimated at high Cobb angles.

## Discussion

There has been attempts to understand the relationship between height loss and Cobb angle for over 50 years^1^. Examples include Bjure et al used a power (log) function to perform regression, Kono and Ylikoski used a linear function^10, 12^, and Stokes et al used a square function^11^. Many different functions have been used, yet the relationship between height loss and Cobb angle has remained unclear. Our novel trigonometric model showed that the Cobb angle could be related to angles in bisected right-angle triangle (ΔBCE in figure 1A) with the AVT representing one side of the tangent of the triangle. We showed a real case application in Figure 8 and our model estimate was better than that by Bjure’s formula.

The new tangent formula showed the least mean bias when applied to both datasets in comparison to the previous regression models. Compared to regression formulae, mathematical models can be widely applied as they are not limited by the dataset used to derive the formula. This can be applied in the clinical setting, as slight differences in surgery may affect extent of curvature correction.

The second advantage of the new Tangent formula is the ability to extrapolate beyond the range of Cobb angle present in the original dataset. The problem of extrapolation is obvious with the power function formula developed by Bjure et al when they did the first attempt to model height loss by Cobb angle^1^. For example, for a patient with 140 degree curve, the Bjure formula predicted a much higher height loss of 23.1 cm when compared with other formulae (tangent formula: 7.6 cm, Kono formula: 8.7 cm, Stokes formula: 17.5 cm). Similar findings of over-estimation was shown in Figure 6. It shows that the ability to extrapolate beyond the range of observed data is limited in regression based models.

Kono formula and Stokes formula required input of more than one Cobb angle values or a decision to choose between single curve or double curve formulae. Kono formula will return very large negative prediction values when many Cobb angles are entered as the formula requires each Cobb angle value to be subtracted by 30 degree. This problem was the cause for large negative bias in Tyrakowski study when using the Kono formula^14^. When applying Kono formula in our local case series, we only included Cobb angles of structural curves into calculation and it gave more reasonable results and a comparable level of bias with other formulae. It illustrates the difficulty and potential confusion if more than one Cobb angle values are required for input into these formulae.

Another unique advantage of the trigonometric model is that it could be used to understand length differences of convex and concave sides of the scoliotic spine which may provide new insight into the differential growth of convex and concave sides of affected vertebrae^16^. Figure 7 demonstrates how to use the trigonometric model to determine the length difference of the convex and concave side of a scoliotic spine.

**Figure 7.**
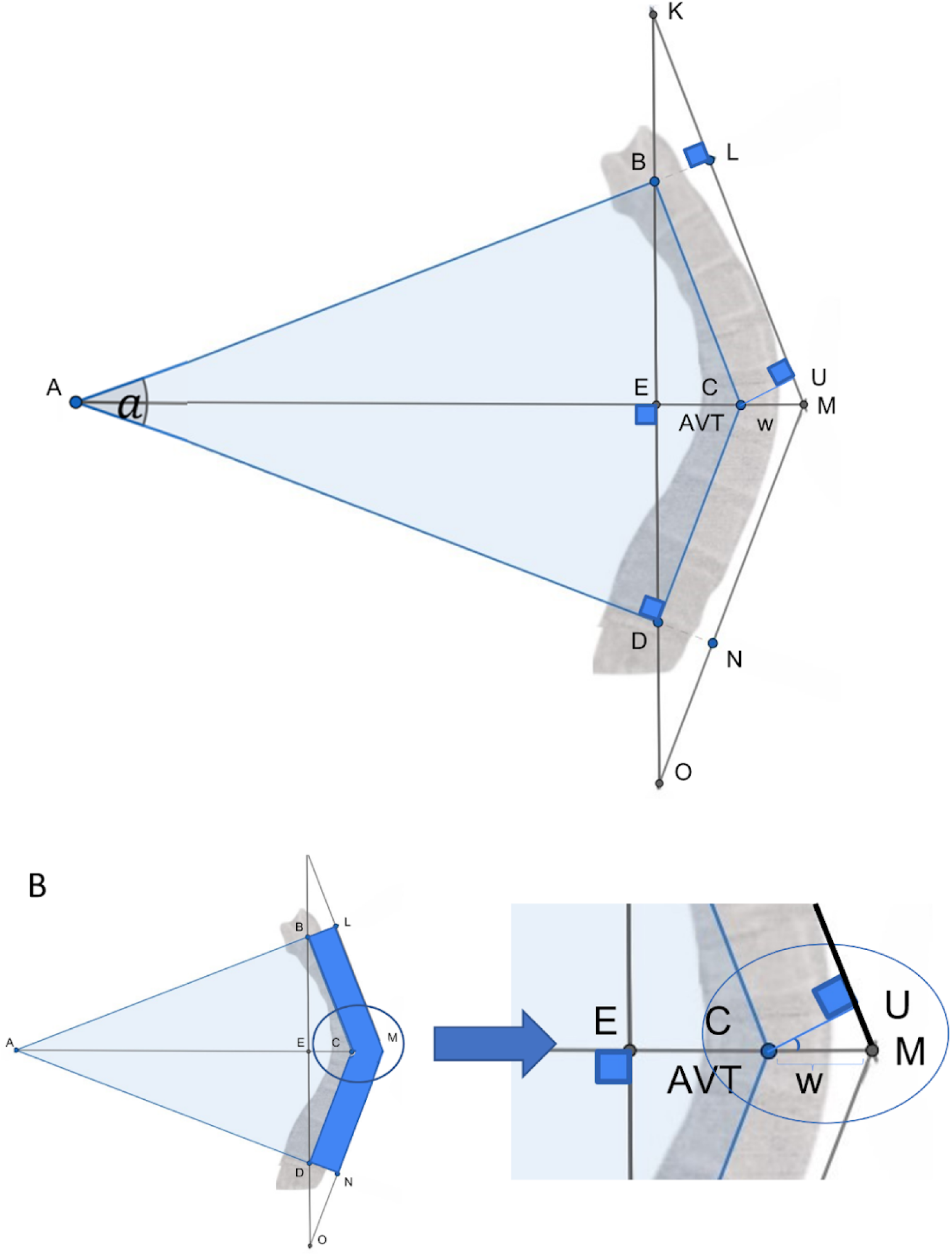
Figure 7A shows the application of the extended Tangent formula on the scoliotic spine with the Concave side = segments BC + CD and Convex side = segment LM + MN. Width of vertebra w is represented by segment CM. ∠MCU = Cobb/2. Figure 7B shows a zoomed in right triangle MCU, the excess in length (ΔH) of the convex side over concave side is represented by 2 times segment UM.

**Figure 8.**
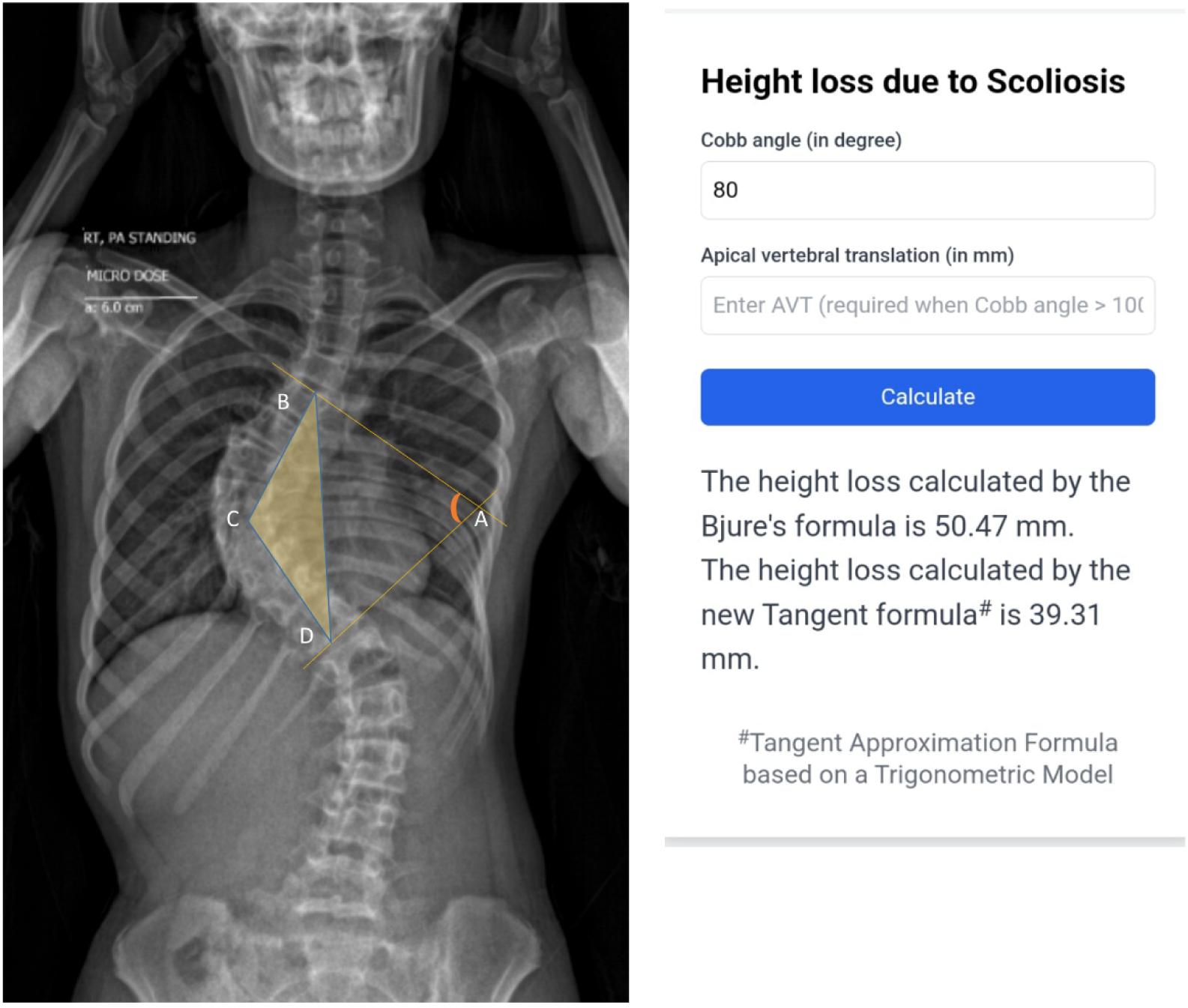
Figure 8 shows application of the model in a prospective patient with a Cobb angle of 80 degree at the major curve. Body height increased by 35 mm (156.5 cm to 160.0 cm). Height loss estimated by this novel trigonometric model was 39 mm while Bjure’s formula returned 50 mm. Labelling of angles in the triangles drawn are similar to those in Figure 1.

Using these two following assumptions:

1. The predicted length of the spine in the Tangent formula represents the length of concave side of affected segment of spine with a given value for vertebra width (w), for example 2.5cm which is the mean mid-vertebral width of T9^17^. Our extended Trigonometric model shows that the length of lines LM + MN is the length of convex side of the involved segment of spine between 2 end vertebrae.
2. This allows the model to derive the length difference is equal to 2 * segment UM. ΔMCU is a right triangle that is ∠MCU = (Cobb/2) and its hypotenuse length = width of the apical vertebra (w). The length difference between the concave and convex lengths of the spine can be solved.

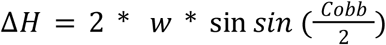

Where Δ*H* = the length difference between concave and convex lengths (cm)

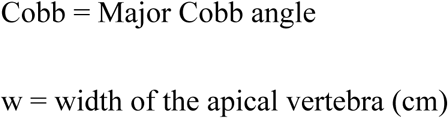

Table 3 shows the predicted length difference of concave and convex lengths calculated with this extended model of scoliosis with a T9 apex for a range of Cobb angles.

**Table 3.**
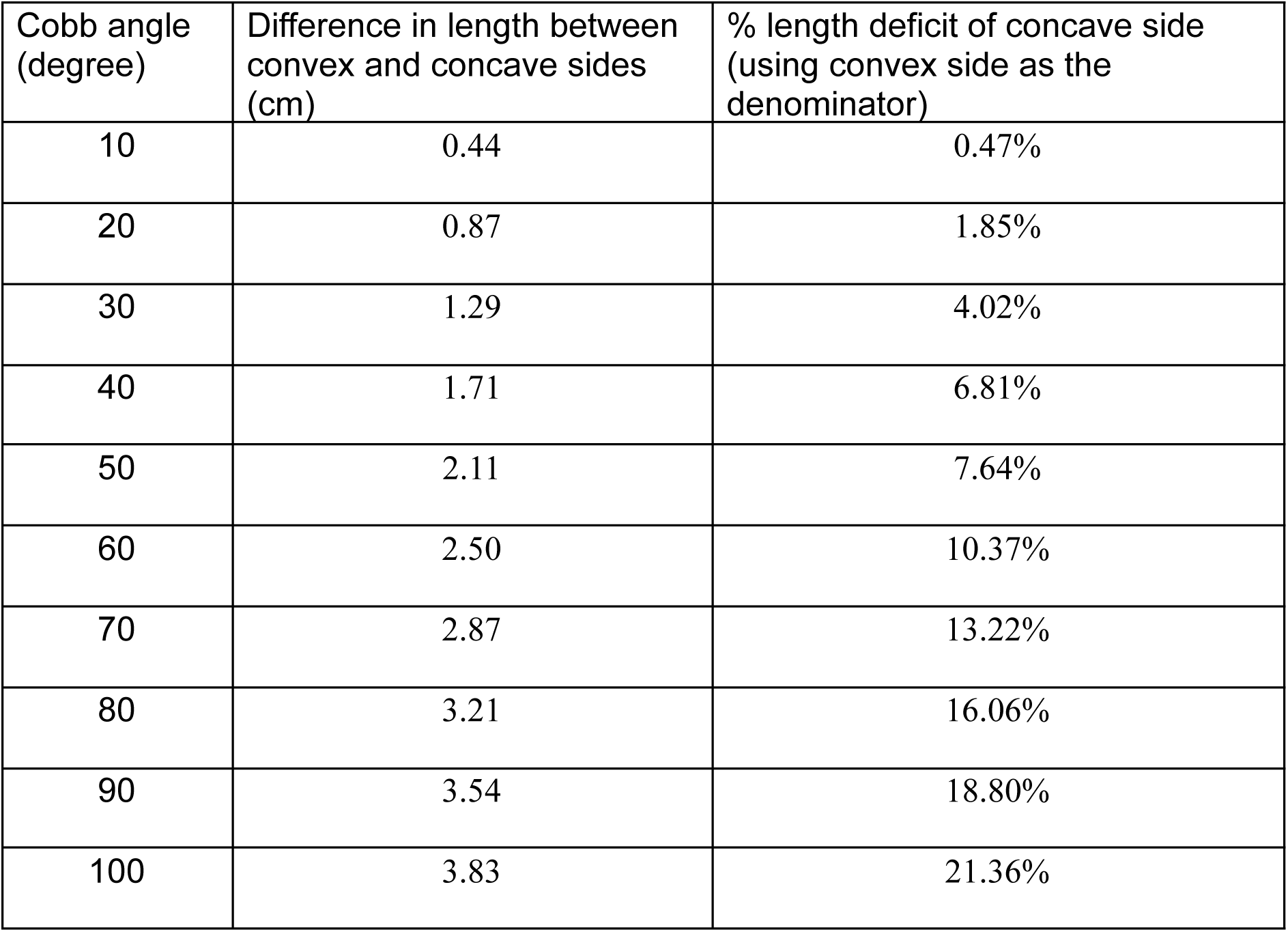
Extended Trigonometric model to determine the differential length of convex and concave sides of scoliosis.

| Cobb angle<br>(degree) | Difference in length between<br>convex and concave sides<br>(cm) | % length deficit of concave side<br>(using convex side as the<br>denominator) |
| --- | --- | --- |
| 10 | 0.44 | 0.47% |
| 20 | 0.87 | 1.85% |
| 30 | 1.29 | 4.02% |
| 40 | 1.71 | 6.81% |
| 50 | 2.11 | 7.64% |
| 60 | 2.50 | 10.37% |
| 70 | 2.87 | 13.22% |
| 80 | 3.21 | 16.06% |
| 90 | 3.54 | 18.80% |
| 100 | 3.83 | 21.36% |

The differential length between 2 sides of vertebrae increases with Cobb angle in a linear manner. On average, the difference increases by ∼0.4 cm for every 10 degree of Cobb angle increase. Table 3 results are comparable to the results measured of CT scans^18^. This length discrepancy can be expressed as % of shortening on the concave. At low Cobb angles (10 to 20 degree), % difference was minimal at below 2%. The intervertebral disc (IVD) to vertebra height ratio is also typically between 1:4 and 1:4.7, such that IVD accounts for 18% to 20% of vertebral body height^19, 20^. The % difference shown in Table 3 also explain the fact that in early scoliosis, loss in height (e.g. below 2% as determined here) is easily accounted by compression of IVD. Furthermore, at Cobb angles of 30 degree and above, more than 4% deficit in length occurs in the concave side and bone wedging may come into play when IVD can no longer accommodate all the difference^21, 22^. This unique feature enables potential stimulation of the complex interaction among asymmetric vertebral growth, spine remodelling and wedging of IVD and vertebrae body in the progression of scoliosis^5, 23, 24^.

### Limitations

In theory, surgical correction is not perfect, and true height is not attained even after operation, but this is a limitation faced by all similar studies. Furthermore, the tangent formula requires the input of AVT. Ideally, AVT is measured for every patient, however it is still not common practice.. Therefore, this model utilises the mean AVT measured by Şarlak et al to ensure that the formula function even without a measured AVT^13^.

In the Şarlak et al dataset, the highest Cobb angle was 75.2 degree. This means there is a lack of data for AVT above this Cobb angle^13^. We also hope that AVT measurement will be more widely practice in the future, so that an individualised calculation could be done for each patient.

Moreover, it is increasingly recognised that spinal deformities and the associated height loss can profoundly affect a patient’s psychological well-being and self-esteem. However, they were not the focus of this study. In the future, quality of life indexes may be used to correlate against predicted height loss. In addition to this, surgical corrections are not perfect in theory. Thus, true height would not be attained even after operation and achieving a return to true height is not feasible.

### A website developed for easy access of the tangent formula

The tangent formula is implemented into this webpage: https://tangentformulae.scoliosisdevices.com/ for easy access. User can simply enter the Cobb angle value (in degree) and it will return the predicted height loss in mm by the tangent formula and Bjure formula. For Cobb angle above 100 degree, it will require user to input of AVT. For lower Cobb angle, the AVT will be assumed following the scheme described here. However, user is also allowed to input AVT and the calculation will take the actual AVT instead of the mean value.

## Conclusion

A new trigonometric concept is used to predict height loss which only requires standard clinical measurements, that is Cobb angle with or without AVT. It shows advantages over previous prediction equations in 2 independent validation patients series and has lower mean bias. The model can be easily extended to understand differential growth or length of convex and concave side of scoliotic spine which provides new insight into the differential growth and extent of wedging in different stages of pathogenesis in scoliosis.

## Data Availability

All data produced in the present work are contained in the manuscript.

https://tangentformulae.scoliosisdevices.com/

## Appendix

### Formulae of estimation of height loss used in this manuscript

#### New Tangent Model Formula

This formula utilizes the trigonometric rules in a triangle modelled after a spine with scoliosis.

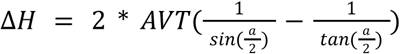

Where AVT = (Apical Vertebral Translation)

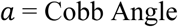

#### Bjure et al. method

This method was developed in a study with 62 scoliotic patients with the age range of 7-25 and was developed for the purpose for increased accuracy in predicting spirometric values i.e vital capacity.

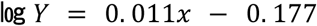

Where Y is the loss of body height caused by Idiopathic Scoliosis (cm)

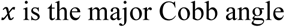

#### Kono et al. method

This method was developed in a study with 140 scoliotic patients with the age range 11-21. Similarly, it also served the purpose of correcting the predictive vital capacity, which is calculated from vital capacity and body height.

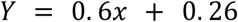

Where Y = the loss of body height caused by Idiopathic Scoliosis (mm)

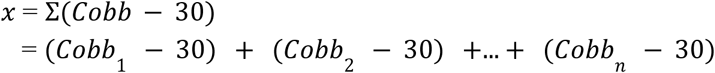

In which each number corresponds to a curve in the spine

#### Stokes et al. method

This method was developed in a study to determine the relationship between Cobb angle, spinal height and spinal length. The study cohort included 407 radiographs and/or patients with the age range of 9-20 prior to any operation.

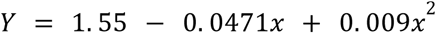

Stokes also developed a second formula for patients with double curves

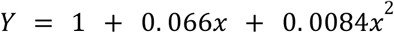

For both formulae:

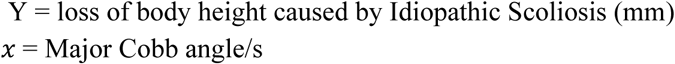

#### Proof of angle CBE = half of Cobb angle (Figure 1)

As the points B and D are points on the endplates of the curvature, and C is the centroid of the apex vertebra. The perpendicular lines to the segments BC and DC intersect to form the Cobb angle. This means that:

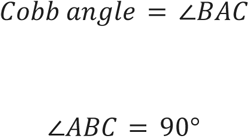

To find the segments BC and BE, the angle CBE can be used. To determine∠*CBE*, the angle ∠*ABE* can be used as the perpendicular lines AB and BC form a right angle. Meaning that:

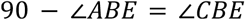

To find∠*ABE*, the law that the sum of angles in a triangle 180° is used, and the angles in the right angled triangle *ABE* can be determined as:

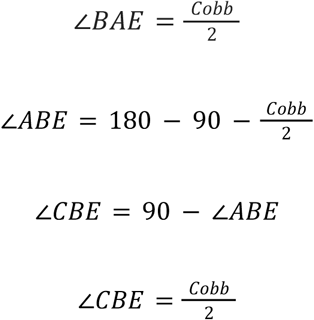

Therefore, ∠CBE in the right triangle BCE (Figure 1C) is equal to half of Cobb angle.

